# Factors Impacting Quality of Life in Multiple System Atrophy

**DOI:** 10.1101/2022.11.06.22281676

**Authors:** Nabila Ali, Vanessa Nesspor, Jee Bang, Sonja W. Scholz, Alexander Pantelyat

## Abstract

**Background:** Multiple system atrophy (MSA) is an atypical parkinsonian disorder that involves parkinsonism, cerebellar ataxia, and autonomic dysfunction. Patient-reported quality of life is an important benchmark for clinicians and clinical trials. The Unified Multiple System Atrophy Rating Scale (UMSARS) allows healthcare providers to rate and assess MSA progression. The MSA-QoL questionnaire is a health-related quality of life scale intended to provide patient-reported outcome measures. In this article, we investigated inter-scale correlations between the MSA-QoL and UMSARS to determine factors impacting the quality of life of patients with MSA.

**Methods:** Twenty patients at the Johns Hopkins Atypical Parkinsonism Center’s Multidisciplinary Clinic with a diagnosis of clinically probable MSA and who filled out the MSA-QoL and UMSARS questionnaires within two weeks of each other were included. Inter-scale correlations between MSA-QoL and UMSARS responses were examined. Linear regressions were also performed to examine relationships between both scales.

**Results:** Significant inter-scale correlations were found between the MSA-QoL and UMSARS, both between MSA-QoL total score and UMSARS Part I subtotal scores and for individual scale items. There were no significant correlations between MSA-QoL life satisfaction rating and UMSARS subtotal scores or any specific UMSARS items. Linear regression analysis found significant associations between MSA-QoL total score and UMSARS Part I and total scores, and between MSA-QoL life satisfaction rating and UMSARS Part I, Part II, and total scores (after adjustment for age).

**Conclusions:** Our study demonstrates significant inter-scale correlations between MSA-QoL and UMSARS, particularly relating to activities of daily living and hygiene. MSA-QoL total score and UMSARS Part I subtotal scores, which assess patients’ functional status, were strongly and significantly correlated. The lack of significant associations between MSA-QoL life satisfaction rating and any UMSARS item suggests there may be aspects to quality of life that are not fully captured by this assessment. Larger cross-sectional and longitudinal analyses utilizing UMSARS and MSA-QoL are warranted and modification of the UMSARS should be considered.

## Introduction

Multiple system atrophy (MSA) is an atypical parkinsonian disorder marked by parkinsonism, cerebellar ataxia, autonomic dysfunction, and a lack of response to dopaminergic medications such as levodopa [1]. The course of MSA is usually aggressive in comparison with that of Parkinson disease, with a median time to wheelchair confinement at five years from disease onset and median time to death at about ten years from disease onset [2], [3]. Clinical manifestations of MSA, including but not limited to motor symptoms, dysphagia, orthostatic hypotension, bladder dysfunction, disordered sleep, and obstructive sleep apnea, significantly impact patient quality of life [1]. Factors associated with a more aggressive disease course include early falls; older age at disease onset; severe autonomic failure; and severe urinary retention [4], [5], [6], [7].

Clinically, MSA is divided into three subtypes, based on symptom predominance: parkinsonian (MSA-P), cerebellar (MSA-C), or mixed. MSA-P presents with parkinsonian signs such as bradykinesia, rigidity, and tremor; MSA-C with signs such as gait and limb ataxia or oculomotor dysfunction; and MSA-mixed with features of both [8]. Though several prior studies found shorter survival time and lower health-related quality of life in MSA-P compared to MSA-C, this was not found to be the case in a large multi-center North American MSA cohort [2], [3], [9], [10]. There are no currently approved disease-modifying MSA treatments, and multiple clinical trials have failed to improve outcome measures, raising the possibility that outcome measures in MSA should be reconsidered. To optimize clinical trial outcomes and to track disease progression in the clinic, it is important to determine the specific patient-reported disease aspects that affect quality of life.

Health-related quality of life (Hr-QoL) is significantly impaired in those with atypical parkinsonian disorders, including MSA [11]. Some specific features of MSA, such as autonomic symptoms (e.g., orthostatic hypotension) and cognitive impairment, have already been demonstrated to be associated with more rapid disease progression and poorer quality of life [12], [13], [14]. MSA is also marked by notable lower urinary tract symptoms, which can require intermittent catheterization and is “a major cause of hospitalization and dependence upon carers” [15]. It is important to understand factors that impact quality of life in individuals with MSA, as it is a potentially modifiable outcome. For this reason, Hr-QoL can serve as a benchmark in assessing disease progression and as a potential clinical trial outcome. To fulfill this purpose, valid and reliable methods of quantifying Hr-QoL and MSA symptom progression are needed.

The MSA-QoL questionnaire was developed as a Hr-QoL scale intended to provide patient-reported outcome measures for this debilitating condition [16]. This scale is the first published and validated patient-reported Hr-QoL measure for patients with MSA. It contains items addressing motor symptoms; nonmotor symptoms such as autonomic dysfunction, sexual impairment, and bowel/bladder dysfunction; and emotional/social impacts of MSA, such as feelings of isolation and anxiety about the future [16]. Patients and/or caregivers are instructed to rate MSA symptoms over a period of 4 weeks, reporting “no problem,” “slight problem,” “moderate problem,” “marked problem,” or “extreme problem” for each symptom in question, with a final item asking patients to report overall life satisfaction from 0 to 100 by marking a grid.

The Unified Multiple System Atrophy Rating Scale (UMSARS) was developed to objectively rate and assess MSA progression [17]. It is commonly used as a primary clinical trial outcome in this condition. UMSARS is composed of four parts: a historical domain (Part I) that assesses motor and autonomic disability based on clinician scoring with patient and/or care partner input, a motor examination domain (Part II), an autonomic examination to assess for orthostatic hypotension (Part III), and a final item rating global disability (Part IV) [17]. Clinicians are asked to rate patient functional status during the preceding 2 weeks for various items from a scale of 0 to 4, with 0 corresponding to normal/unimpaired function and 4 corresponding to severe impairment. The clinician also assigns a global disability rating from 1 to 5, with a rating of 1 corresponding to complete independence and a rating of 5 to a totally dependent and bedridden state.

The aim of this cross-sectional study was to specifically explore inter-scale correlations between the MSA-QoL items and subscales and UMSARS items and subscales. We aimed to investigate areas of concordance and discordance between patient-reported and clinician-scored responses, providing insight into factors affecting quality of life in individuals with MSA. Previous work by Meissner *et al*. found that MSA-QoL scores were less indicative of disease progression over time than UMSARS scores [18]. Expanding upon this work, this analysis also explored which individual patient-reported outcomes correlated with clinician-scored disease severity, impact, and functional status.

## Methods

Twenty patients who were assessed at the Johns Hopkins Atypical Parkinsonism Center’s Multidisciplinary Clinic between 2015 and 2022 by one of the study co-authors (J.B., S.W.S., or A.P.) were included in this study. This study received Johns Hopkins Institutional Review Board approval (IRB00062534). Study inclusion criteria were: (1) clinically probable MSA diagnosis (retrospectively determined according to the recently published MSA diagnostic criteria; “MSA-mixed” designation was assigned when patients had features of both parkinsonism and cerebellar dysfunction without clear predominance of one over the other) and (2) availability of MSA-QoL and UMSARS questionnaires completed within two weeks of each other [8]. The UMSARS was administered during the clinic visit and the MSA-QoL were mailed (or emailed, as per patient preference) to patients to complete and return.

### MSA-QoL Questionnaire

The MSA-QoL is a 40-item questionnaire that asks patients to rate their level of difficulty with specific mental or physical domains, as well as a question asking patients to mark overall life satisfaction on a scale from 0 to 100. It was developed based on physician and patient reports and psychometric data analysis and has been shown to be reliable and valid [16]. Items include questions about mobility, coordination, ability to complete self-care, other activities of daily living, symptoms of dysautonomia, sleep quality, cognitive function, and social/emotional impacts of the disease. In total, there are 14 motor items, 12 nonmotor items, and 14 emotional/social items. Higher total scores on the MSA-QoL indicate higher levels of impairment, and higher scores on the life satisfaction item show higher life satisfaction.

### UMSARS

UMSARS is a clinician-scored rating scale to assess the severity of symptoms for patients with MSA. There is a historical component (Part I) with 12 items, a motor examination component (Part II) with 14 items, an optional autonomic examination component (Part III), and a global disability scale (Part IV). UMSARS has also been demonstrated to be reliable and valid [17]. In the UMSARS, symptoms are rated over a period of two weeks. Higher UMSARS scores indicate greater levels of impairment.

### Statistical Analysis

The Kruskal-Wallis rank-sum test or Pearson’s χ^2^ test was used to compare patient characteristics between groups. Pearson’s correlations with Benjamini-Hochberg correction (to adjust for multiple comparisons) between MSA-QoL items and UMSARS items were calculated to examine inter-scale associations [19]. Linear regression models using MSA-QoL scores as the outcome variable and UMSARS scores as the predictor variable, adjusting for age, gender, disease duration, and disease type, were used to examine the relationships between both scales. Correlation coefficients with *p* < 0.05 were considered to be statistically significant. All statistical analyses were performed in RStudio version 2021.09.2+382 (RStudio, PBC, 2022).

## Results

Patient characteristics are summarized in Table 1. There were no significant differences in age, sex, or disease duration among MSA types (MSA-C, MSA-P, or MSA-mixed). There were also no significant differences in MSA-QoL total score, MSA-QoL satisfaction rating, UMSARS Part I subtotal score, UMSARS Part II subtotal score, or UMSARS global disability score among MSA types.

**Table 1.** Patient characteristics.

| Characteristic | All patients | MSA-C | MSA-P | MSA-mixed | Adjusted p-value |
| --- | --- | --- | --- | --- | --- |
| *N* | 20 | 5 | 12 | 3 |  |
| Age in years, mean (SD) | 61.3 (7.1) | 61.8 (7.3) | 61.8 (8.0) | 58.0 (3.5) | 0.4307 |
| Sex, *n* (%) |  |  |  |  | 0.7659 |
| Female | 10 (50%) | 3 (60%) | 6 (50%) | 1 (33%) |  |
| Male | 10 (50%) | 2 (40%) | 6 (50%) | 2 (67%) |  |
| Disease duration in years, mean (SD) | 5.3 (3.1) | 4.6 (2.7) | 5.3 (2.9) | 6.3 (4.9) | 0.8270 |
| MSA-QoL |  |  |  |  |  |
| Total, mean (SD) | 86.6 (29.9) | 69.0 (29.8) | 93.1 (30.3) | 92.0 (25.2) | 0.3058 |
| Satisfaction, mean (SD) | 49% (23) | 62% (16) | 48% (24) | 29% (19) | 0.0868 |
| UMSARS |  |  |  |  |  |
| Part I subtotal score, mean (SD) | 27.8 (7.8) | 24.6 (5.3) | 27.9 (9.0) | 32.3 (4.2) | 0.2089 |
| Part II subtotal score, mean (SD) | 30.2 (10.1) | 27.4 (11.7) | 30.3 (10.7) | 34.7 (2.5) | 0.5506 |
| Total score (SD) | 58.0 (17.0) | 52.0 (15.7) | 58.2 (19.0) | 67.0 (6.1) | 0.3446 |
| Global disability score, mean (SD) | 3.3 (1.1) | 2.8 (1.3) | 3.3 (1.1) | 4.0 (0.0) | 0.2900 |

We discovered significant inter-scale correlations between subtotal scores on individual MSA-QoL items and UMSARS Part I (History), UMSARS Part II (Motor Examination), UMSARS total score, UMSARS global disability rating; all *p*-values reported below are adjusted for multiple comparisons. The findings are described in Figure 1 and in the text below.

**Figure 1.**
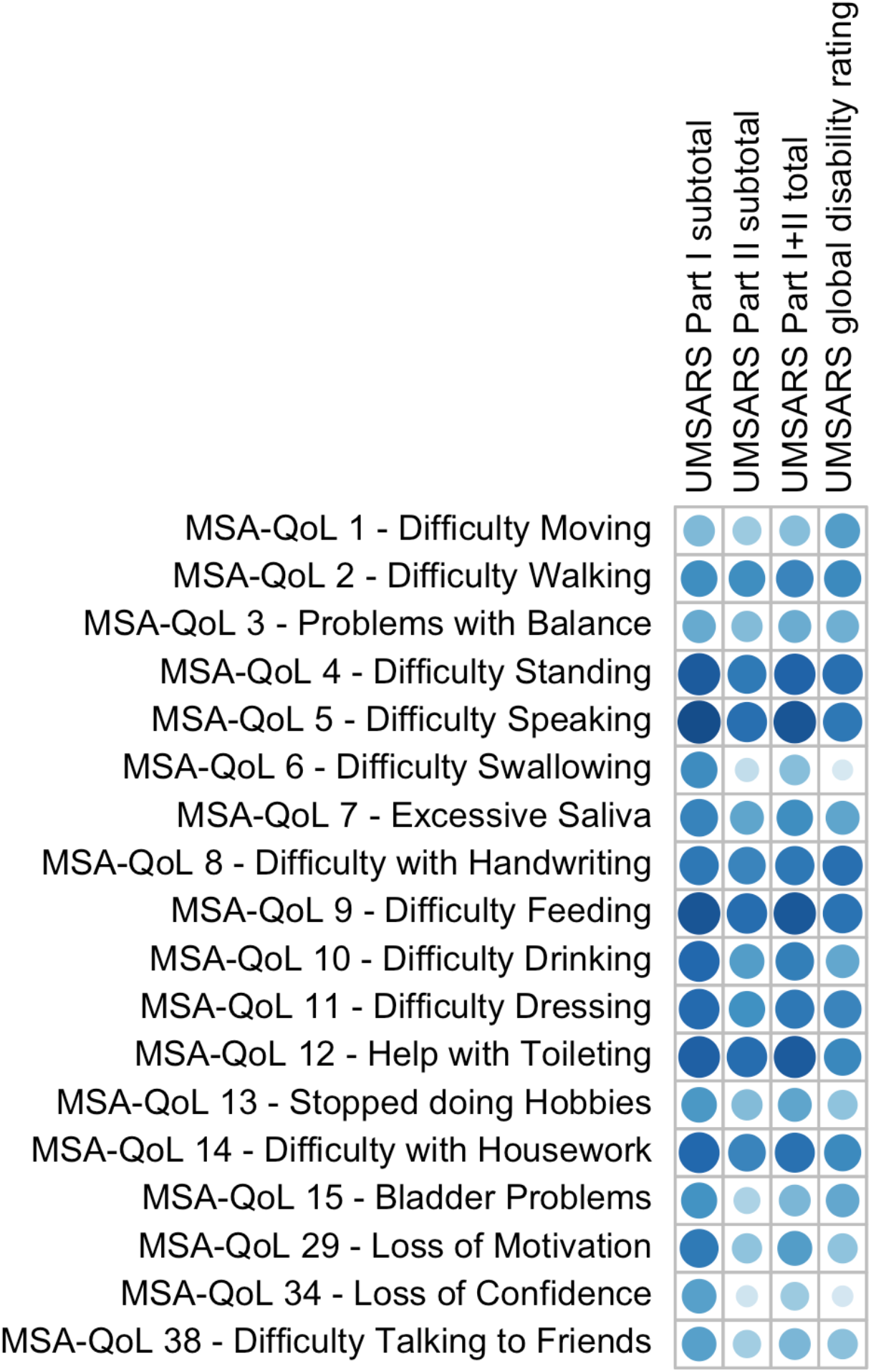
A visual representation of the correlations between UMSARS domains and selected individual MSA-QoL items. A larger and darker circle represents a correlation coefficient that is closer to 1.

**Figure 2.**
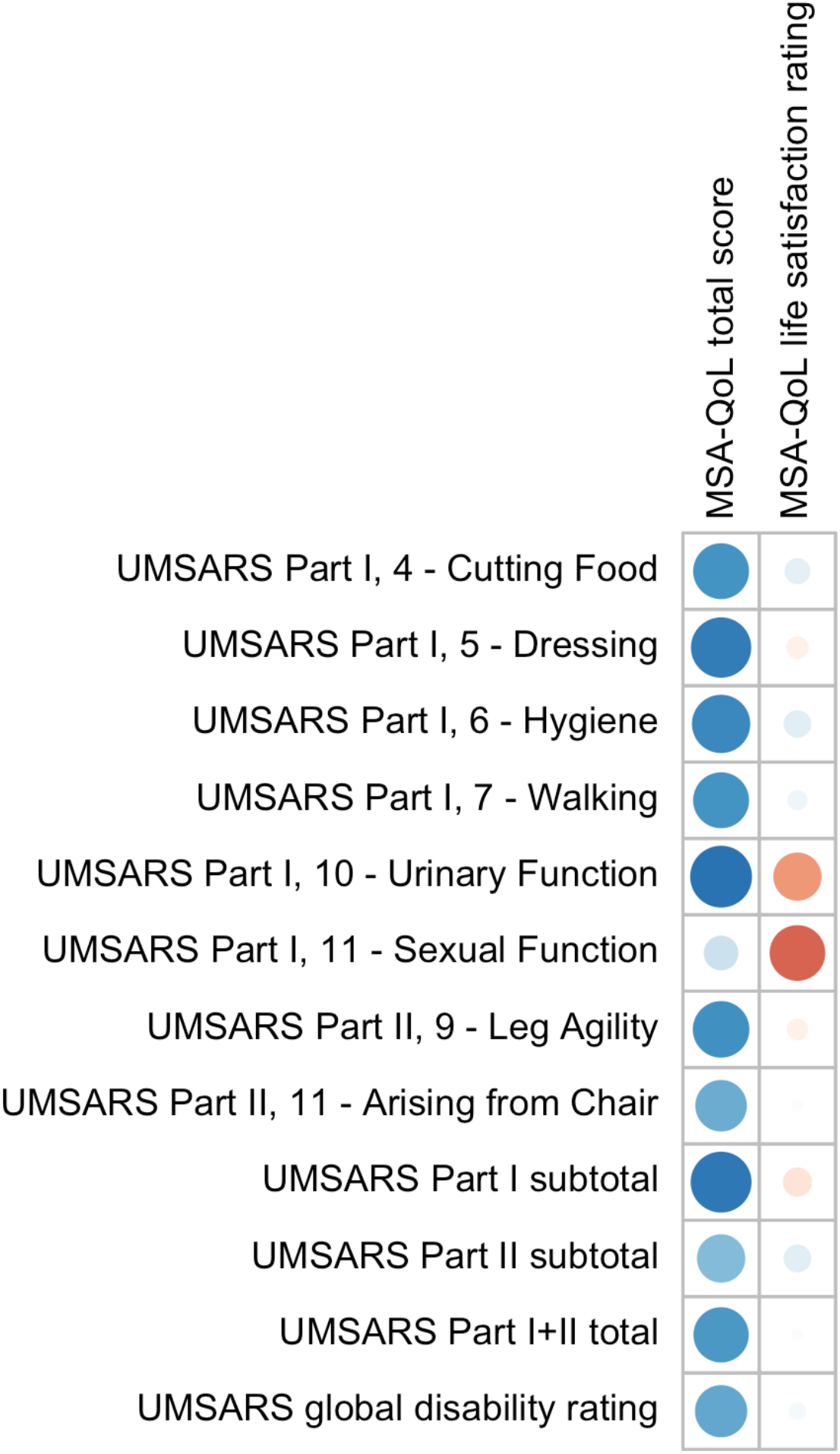
A visual representation of the correlations between MSA-QoL domains and selected individual UMSARS items. A large blue circle represents a correlation coefficient that is close to 1, and a large red circle represents a correlation coefficient that is close to -1.

Specifically, UMSARS Part I subtotal scores were significantly correlated with MSA-QoL items 4—Difficulty Standing (*r* = 0.8394, adjusted *p* = 0.0014), 5—Difficulty Speaking (*r* = 0.8812, *p* = 0.0004), 7—Excessive Saliva (*r* = 0.6603, *p* = 0.0326), 8—Difficulty with Handwriting (*r* = 0.7156, *p* = 0.0153), 9—Difficulty Feeding (*r* = 0.8502, *p* = 0.0010), 10— Difficulty Drinking (*r* = 0.7852, *p* = 0.0052), 11—Difficulty Dressing (*r* = 0.7736, *p* = 0.0060), 12—Help with Toileting (*r* = 0.8154, *p* = 0.0026), 14—Difficulty with Housework (*r* = 0.7819, *p* = 0.0053), and 29—Loss of Motivation (*r* = 0.7001, *p* = 0.0196). UMSARS Part I subtotal scores were also correlated with MSA-QoL items 2—Difficulty Walking (*r* = 0.6169, *p* = 0.0539), 3— Problems with Balance (*r* = 0.5039, *p* = 0.1511), 6—Difficulty Swallowing (*r* = 0.6205, *p* = 0.0518), 13—Stop Doing Hobbies (*r* = 0.5775, *p* = 0.0825), 15—Bladder Problems (*r* = 0.5988, *p* = 0.0654), 34—Loss of Confidence (*r* = 0.5467, *p* = 0.1083), and 38—Difficulty Talking to Friends (*r* = 0.5444, *p* = 0.1105), but these correlations were not significant after adjustment for multiple comparisons.

UMSARS Part II subtotal scores were significantly correlated with MSA-QoL items 4— Difficulty Standing (*r* = 0.7045, *p* = 0.0184), 5—Difficulty Speaking (*r* = 0.7561, *p* = 0.0080), 8—Difficulty with Handwriting (*r* = 0.6524, *p* = 0.0359), 9—Difficulty Feeding (*r* = 0.7660, *p* = 0.0067), 12—Help with Toileting (*r* = 0.7663, *p* = 0.0067), and 14—Difficulty with Housework (*r* = 0.6566, *p* = 0.0341). UMSARS Part II subtotal scores were also correlated with MSA-QoL items 2—Difficulty Walking (*r* = 0.6151, *p* = 0.0549), 7—Excessive Saliva (*r* = 0.5218, *p* = 0.1321), 10—Difficulty Drinking (*r* = 0.5575, *p* = 0.1004), and 11—Difficulty Dressing (*r* = 0.6052, *p* = 0.0606), but these correlations were not significant after adjustment for multiple comparisons.

UMSARS combined Part I and Part II scores were significantly correlated with MSA-QoL items 2—Difficulty Walking (*r* = 0.6506, *p* = 0.0366), 4—Difficulty Standing (*r* = 0.8022, *p* = 0.0036), 5—Difficulty Speaking (*r* = 0.8526, *p* = 0.0010), 8—Difficulty with Handwriting (*r* = 0.7164, *p* = 0.0151), 9—Difficulty Feeding (*r* = 0.8454, *p* = 0.0012), 10—Difficulty Drinking (*r* = 0.6868, *p* = 0.0232), 11—Difficulty Dressing (*r* = 0.7117, *p* = 0.0161), 12—Help with Toileting (*r* = 0.8306, *p* = 0.0018), and 14—Difficulty with Housework (*r* = 0.7475, *p* = 0.0089). UMSARS combined Part I and Part II scores were also correlated with MSA-QoL items 3—Problems with Balance (*r* = 0.4904, *p* = 0.1678), 6—Difficulty Swallowing (*r* = 0.4213, *p* = 0.2495), 13—Stopped doing Hobbies (*r* = 0.5206, *p* = 0.1334), and 29—Loss of Motivation (*r* = 0.5552, *p* = 0.1019), but these correlations were not significant after adjustment for multiple comparisons.

UMSARS global disability scores were significantly correlated with individual MSA-QoL items 2—Difficulty Walking (*r* = 0.6377, *p* = 0.0425), 4—Difficulty Standing (*r* = 0.7589, *p* = 0.0077), 5—Difficulty Speaking (*r* = 0.7174, *p* = 0.0148), 8—Difficulty with Handwriting (*r* = 0.7519, *p* = 0.0085), 9—Difficulty Feeding (*r* = 0.7322, *p* = 0.0119), 11—Difficulty Dressing (*r* = 0.6590, *p* = 0.0332), 12—Help with Toileting (*r* = 0.6485, *p* = 0.0373), and 14—Difficulty with Housework (*r* = 0.6367, *p* = 0.0429). UMSARS global disability scores were also correlated with MSA-QoL items 1—Difficulty Moving (*r* = 0.5576, *p* = 0.1004), 3—Problems with Balance (*r* = 0.4859, *p* = 0.1737), 7—Excessive Saliva (*r* = 0.5238, *p* = 0.1302), 10—Difficulty Drinking (*r* = 0.5101, *p* = 0.1450), and 15—Bladder Problems (*r* = 0.5105, *p* = 0.1448), but these correlations were not significant after adjustment for multiple comparisons.

Corresponding individual items on UMSARS and MSA-QoL also agreed well with each other, though there were some notable discrepancies. These findings are described in Table 2 below.

**Table 2.** Correlations of individual UMSARS items with closest corresponding individual MSA-QoL items. Significant correlations (p < 0.05) after adjusting for multiple comparisons are bolded.

| UMSARS Item | MSA-QoL Item | $r$ (adjusted $p$) |
| --- | --- | --- |
| UMSARS Part I, 7—Walking | MSA-QoL 2—Difficulty Walking | 0.6657 (**0.0301**) |
| UMSARS Part I, 8—Falling | MSA-QoL 3—Problems with Balance | 0.0365 (0.9511) |
| UMSARS Part I, 1—Speech | MSA-QoL 5—Difficulty Speaking | 0.7298 (**0.0123**) |
| UMSARS Part I, 2—Swallowing | MSA-QoL 6—Difficulty Swallowing | 0.6055 (0.0606) |
| UMSARS Part I, 3—Handwriting | MSA-QoL 8—Difficulty with Handwriting | 0.6496 (**0.0368**) |
| UMSARS Part I, 4—Cutting Food | MSA-QoL 9—Difficulty Feeding | 0.8221 (**0.0022**) |
| UMSARS Part I, 5—Dressing | MSA-QoL 11—Difficulty Dressing | 0.8699 (**0.0006**) |
| UMSARS Part I, 10—Urinary Function | MSA-QoL 15—Bladder Problems | 0.7930 (**0.0042**) |
| UMSARS Part I, 12—Bowel Function | MSA-QoL 16—Constipation | 0.3069 (0.4476) |
| UMSARS Part I, 9—Orthostatic Symptoms | MSA-QoL 17—Dizziness when Standing | 0.3052 (0.4490) |

MSA-QoL total score was significantly correlated with UMSARS Dressing (*r* = 0.6950, *p* = 0.0207), UMSARS Hygiene (*r* = 0.6479, *p* = 0.0375), and UMSARS Urinary Function (*r* = 0.7332, *p* = 0.0117). There were significant correlations between MSA-QoL total score and UMSARS Cutting Food (*r* = 0.5914, *p* = 0.0715), UMSARS Walking (*r* = 0.5928, *p* = 0.0706), UMSARS Leg Agility (*r* = 0.6077, *p* = 0.0594), and UMSARS Arising from Chair (*r* = 0.4987, *p* = 0.1572), but these correlations were no longer significant after adjustment for multiple comparisons. MSA-QoL total score and UMSARS Part I subtotal score were strongly and significantly correlated (*r* = 0.7101, *p* = 0.0165). MSA-QoL total score and UMSARS total score, obtained from summing UMSARS Part I and Part II subtotal scores, were significantly correlated (*r* = 0.5785, *p* = 0.0818), as were MSA-QoL total score and UMSARS global disability rating (*r* = 0.5129, *p* = 0.1421), but this correlation was no longer significant after adjustment for multiple comparisons. MSA-QoL total score and UMSARS Part II subtotal score were not significantly correlated (*r* = 0.4361, *p* = 0.2323).

There was a moderate negative correlation between MSA-QoL life satisfaction rating and UMSARS Sexual Function (*r* = -0.5878, *p* = 0.0740), but this correlation was not significant after adjustment for multiple comparisons. There was a strong and significant correlation between MSA-QoL life satisfaction and MSA-QoL item 17—Dizziness when Standing (*r* = -0.7281, *p* = 0.0125). There were no other significant correlations between MSA-QoL life satisfaction rating and any other MSA-QoL individual items or any UMSARS individual items or subtotal scores, including UMSARS Part I total (*r* = -0.1451, *p* = 0.7600), UMSARS Part II total (*r* = 0.1287, *p* = 0.7868), UMSARS total score (*r* = 0.0183, *p* = 0.9767), and UMSARS global disability score (*r* = 0.0437, *p* = 0.9373).

In an exploratory analysis, we also considered associations between the MSA-QoL and UMSARS questionnaires using linear regression models, as shown in Tables 3 and 4. MSA-QoL total scores were not significantly associated with age, sex, disease duration, or MSA type alone (all *p* > 0.32). Parsimony was assessed using the Akaike Information Criterion, which showed that the addition of the covariates age, sex, disease duration, and MSA type did not improve the parsimony of the models [20], [21]. MSA-QoL total score was significantly associated with UMSARS Part I scores (*p* = 0.0058 without adjusting for age, sex, disease duration, or MSA type) and UMSARS combined Part I and II scores (*p* = 0.0170) and tended to be associated with UMSARS Part II scores (*p* = 0.0590) and UMSARS global disability rating (*p* = 0.0563).

**Table 3.** Linear regression model results for MSA-QoL total score, not adjusted for age, sex, disease duration, or MSA type.

|  | MSA-QoL predictors |  |  |  |
| --- | --- | --- | --- | --- |
|  | UMSARS Part I | UMSARS Part II | UMSARS Part I + Part II total | UMSARS global disability |
| Beta (SE) | 2.278 (0.723) | 1.2784 (0.6317) | 0.9292 (0.3513) | 12.133 (5.924) |
| *p*-value | **0.0058** | 0.0590 | **0.0170** | 0.0563 |

**Table 4.** Linear regression model results for MSA-QoL life satisfaction rating, with age as a covariate.

|  | MSA-QoL predictors |  |  |  |
| --- | --- | --- | --- | --- |
|  | UMSARS Part I | UMSARS Part II | UMSARS Part I + Part II total | UMSARS global disability |
| Beta (SE) | -0.004043<br>(0.006526) | -0.003242<br>(0.005656) | -0.002019<br>(0.003203) | -0.010472<br>(0.046319) |
| *p*-value | 0.5443 | 0.5744 | 0.5374 | 0.8240 |
| F-statistic *p*-value | **0.0456** | **0.0468** | **0.0453** | 0.0537 |

MSA-QoL life satisfaction ratings were significantly associated with age alone (*b* = 0.0182, *p* = 0.0144) and sex alone (b = 0.2087, *p* = 0.0463), with female sex associated with higher life satisfaction rating. MSA-QoL life satisfaction ratings were not significantly associated with disease duration or MSA type. After adjustment for age, MSA-QoL life satisfaction was significantly associated with UMSARS Part I scores (*p* = 0.0456 after adjustment for age), UMSARS Part II scores (*p* = 0.0468), and UMSARS combined Part I and II scores (*p* = 0.0453).

## Discussion

This study found strong inter-scale correlations between UMSARS subscale scores and individual MSA-QoL items. MSA-QoL items relating to activities of daily living, such as feeding oneself, dressing oneself, and toileting, were significantly correlated with UMSARS Part I (History) subtotal scores. MSA-QoL items relating to difficulty standing, speaking, drinking, and handwriting, as well as excessive salivation and difficulty with housework, were also significantly correlated with UMSARS Part I subtotal scores. These correlations were strong (*r* = 0.6603 – 0.8812). Many of these MSA-QoL items were also significantly correlated with UMSARS Part II (Motor Examination) subtotal scores, including: difficulty standing, difficulty speaking, difficulty feeding, difficulty with handwriting, requiring help with toileting, and difficulty with housework. These correlations were also strong (*r* = 0.6524 – 0.7663).

UMSARS total scores, obtained by summing UMSARS Part I and Part II scores, were significantly correlated (*r* = 0.6506 – 0.8526) with nine individual MSA-QoL items: difficulty walking, difficulty standing, difficulty speaking, difficulty with handwriting, difficulty feeding, difficulty drinking, difficulty dressing, requiring help with toileting, and difficulty with housework. UMSARS global disability ratings were strongly and significantly correlated (*r* = 0.6367 – 0.7589) with eight individual MSA-QoL items: difficulty walking, difficulty standing, difficulty speaking, difficulty with handwriting, difficulty feeding, difficulty dressing, requiring help with toileting, and difficulty with housework. These findings suggest that disease severity, as assessed by the UMSARS total score and by the clinician assessment of UMSARS global disability, is associated with the ability to complete activities of daily living and other activities related to independence and dignity. Autonomic symptoms, such as excessive salivation, may also be markers of disease severity.

We found no significant correlations between MSA-QoL life satisfaction rating and any UMSARS item, or with any other MSA-QoL item other than 17—Dizziness when Standing (*r* = -0.7281, *p* = 0.0125). This suggests that orthostatic symptoms may be an important contributor to or correlate of life satisfaction, but also that there are aspects to patient satisfaction with their quality of life that are not fully captured by these assessments and may be determined by external factors. These factors may include degree of care partner support, resilience, and levels of optimism/pessimism.

MSA-QoL total scores, obtained from summing all patient-reported responses to questionnaire items, were strongly and significantly correlated to UMSARS Part I subtotal scores (*r* = 0.7101, *p* = 0.0165). This is likely because both domains assess patient functional status, albeit in differing ways: the MSA-QoL assesses patient functional status through patient/caregiver self-report, while UMSARS involves clinician interview of the patient/caregiver. Notably, MSA-QoL total score was also significantly associated with the following UMSARS individual items: dressing, hygiene, and urinary function. This suggests that hygiene issues, as well as issues related to urinary incontinence (likely impacting patients’ sense of dignity as well as hygiene), and independence related to dressing, may be of particular importance to determination of quality of life in those with MSA.

While most individual items on UMSARS that had corresponding MSA-QoL counterparts correlated well (*r* = 0.6496 – 0.8699) with each other, there were some notable exceptions, especially for the items related to falling, dizziness/orthostatic symptoms, and bowel function/constipation. The lack of expected findings may be due to differences in how both questionnaires are administered and interpreted. This is because the MSA-QoL is filled out by the patient and/or caregiver, whereas UMSARS ratings are determined from clinicians’ perceptions of patient responses, adding an additional interpretive layer. Importantly, the MSA-QoL questionnaire also instructs patients to rate symptoms over a period of four weeks, whereas UMSARS instructs providers to rate symptoms over a period of two weeks. Finally, while certain items on the MSA-QoL questionnaire and the UMSARS may pertain to similar domains, subtle differences in the phrasing of corresponding inter-scale items may lead to different responses. For example, MSA-QoL item 17—Dizziness when Standing asks patients to rate severity of dizziness while standing up, while the corresponding UMSARS item asks about “orthostatic symptoms” more generally, including dizziness but also syncope, visual disturbances, and neck pain.

Limitations of this study include the small sample size, its cross-sectional nature, and lack of autopsy confirmation for MSA diagnosis. The small sample size restricted our ability to detect significant differences in our multivariate linear models. The cross-sectional nature of our study also meant we could not examine whether changes in UMSARS scores could predict changes in MSA-QoL over time. We emphasize that this is an exploratory analysis and recognize that the sample size limits the ability to generalize from our findings. However, the reported results provide useful information to inform future larger longitudinal studies.

To summarize, our study demonstrates strong inter-scale correlations between multiple UMSARS items and MSA-QoL items. Particularly, MSA-QoL items relating to activities of daily living, hygiene, and other basic functions were found to have strong and significant correlations with MSA disease severity as determined from UMSARS Part I, Part II, and combined Part I and II scores. This is consistent with other work, which has demonstrated that autonomic symptoms are associated with more rapid disease progression in MSA [12]. Clinicians and clinical researchers should consider the importance of these outcomes when assessing quality of life in patients with MSA. In particular, hygiene and dignity issues, especially those related to urinary incontinence, may be an important and highly specific benchmark when considering quality of life and symptom severity in patients with MSA, as has already been suggested [15].

These results also suggest the possibility of creating a more focused assessment that may accurately capture important aspects of key symptom severity and impact on quality of life. While the full MSA-QoL questionnaire provides valuable information about patient quality of life, the entire survey may be too burdensome and time-consuming to complete during a time-limited healthcare visit. By focusing on activities of daily living and hygiene, which correlate strongly to UMSARS items, clinicians can guide history-taking to focus on patient-reported outcomes that correlate most strongly with clinician-scored outcomes. We also suggest that there may be aspects to overall quality of life that are not fully captured by the UMSARS and that a revision of this assessment should be considered. Although small and cross-sectional in nature, our study draws clear associations between clinician-administered and patient-reported outcomes relevant to quality of life in MSA.

## Data Availability

All data produced in the present study are available upon reasonable request to the authors.

## Statement of Ethics

This study protocol was approved by the Institutional Review Board at Johns Hopkins School of Medicine (Johns Hopkins IRB-2, study number IRB00062534). All participants provided written informed consent for this study.

## Acknowledgments

The authors would like to thank the patients and their families for contributing to this research study. SWS was supported by the Intramural Research Program of the National Institute of Neurological Disorders and Stroke, National Institutes of Health (project number ZIANS003154).

