## Supplemental Table 1 for "Factors Impacting Quality of Life in Multiple System Atrophy"

**Supplementary Information**

***Table S1.*** *Relevant MSA-QoL and UMSARS Part I questionnaire items. Asterisked MSA-QoL items are those for which a statistically significant correlation (after correction for multiple comparisons) was found with UMSARS subscale score(s).*

| MSA-QoL Item | Question Text | UMSARS Item | Question Text |
|---|---|---|---|
| 2 | Had difficulty walking? | Part I, 7 | Walking |
| 3 | Had problems with your balance? | Part I, 8 | Falling (rate the past month) |
| 4 | Had difficulty standing up without support? | | |
| 5 | Had difficulty speaking? | Part I, 1 | Speech |
| 6 | Had difficulty swallowing food? | Part I, 2 | Swallowing |
| 7 | Had too much saliva or drooling? | | |
| 8 | Had difficulty with handwriting? | Part I, 3 | Handwriting |
| 9 | Had difficulty feeding yourself? | Part I, 4 | Cutting food and handling utensils |
| 10 | Had difficulty drinking fluids? | | |
| 11 | Had difficulty dressing yourself? | Part I, 5 | Dressing |
| 12 | Needed help to go to the toilet? | | |
| 13 | Had to stop doing things that you liked to do, e.g., your hobbies? | | |
| 14 | Had difficulty doing things around the house, e.g., housework? | | |
| 15 | Experienced bladder problems? | Part I, 10 | Urinary function |
| 16 | Experienced problems with constipation? | Part I, 12 | Bowel function |
| 17 | Experienced dizziness when standing up? | Part I, 9 | Orthostatic symptoms |
| 23 | Been feeling tired very quickly (without exerting yourself)? | | |
| 38 | Had difficulty talking to friends about your illness? | | |
| | | Part I, 6 | Hygiene |
| | | Part II, 1 | Facial Expression |
| | | Part II, 9 | Leg Agility |
| | | Part II, 11 | Arising from Chair |
| | | Part II, 12 | Posture |
